# Validation of the Qualisex Questionnaire for Psychometric Assessment of Sexual Health in Lichen Sclerosus

**DOI:** 10.64898/2026.10.16.26365725

**Authors:** Tonia Samela, Roberta Priori, Laura Colonna, Lavinia Zozi, Valentina Gallo, Chiara Assorgi, Simone Garcovich, Alessia Paganelli

## Abstract

**Introduction:** Lichen sclerosus (LS) is a chronic inflammatory dermatosis predominantly affecting the anogenital region of both women and men. Sexual dysfunction remains among the most debilitating and overlooked consequences of the disease, as available patient-reported outcome measures are either restricted to one sex or capture sexual health only partially. The aim of the present work is to validate the Qualisex questionnaire (in its the Italian version) in patients with LS and to evaluate its psychometric properties, factor structure, convergent, discriminant validity, and sex symmetry. Methods: A cross-sectional study was conducted on 211 patients (167 women, 44 men; mean age 52.9 years) with a confirmed diagnosis of LS. Participants completed the Italian Qualisex, Skindex-29, DASS-21, and sexual-health-specific scales, the Female Sexual Function Index (FSFI) for women, and the International Index of Erectile Function (IIEF) for men. Construct validity was assessed via Principal Component Analysis (PCA), and reliability via Cronbach’s α. Convergent and discriminant validities were evaluated against standard psychometric batteries.

**Results:** Item 2 was removed due to low communality (0.167). PCA on the remaining 9 items revealed a two-factor structure accounting for 72.5% of total variance: Component 1 (“Sexual Functioning and Satisfaction”, 59.89%) and Component 2 (“Partner Devaluation”, 12.66%). Internal consistency was high (α= 0.911 overall; α= 0.841 in women; α= 0.899 in men). Qualisex scores correlated strongly with Skindex-29 Social Functioning (r = 0.582) and Emotions (r = 0.531) subscales, as well as FSFI (r = −0.391) and IIEF (ρ = −0.566). Discriminant validity was supported by low-to-moderate correlations with DASS-21 domains (r < 0.38). While FSFI and IIEF showed significant negative correlations with age (p = 0.002), Qualisex remained age-independent across sexes (p > 0.05).

**Conclusion:** The 9-item Italian Qualisex is a reliable, sex-symmetrical, and age-independent patient-reported outcome measure for assessing sexual health impairment in LS, successfully isolating disease-specific distress from physiological aging and general distress.

## Introduction

Lichen sclerosus (LS) is a chronic, inflammatory dermatosis that predominantly affects the anogenital region, although extragenital involvement is also described. Its pathogenesis is not fully understood, but autoimmune, genetic, and local factors are all thought to play a role, and the condition typically follows a relapsing-remitting course requiring long-term management[1–3]. While LS has traditionally been considered a disease of postmenopausal women, this view has been increasingly challenged: growing evidence indicates that penile and perianal LS in men are far from rare, and are likely underrecognized due to underreporting, delayed diagnosis, and a persistent lack of awareness among both patients and clinicians[4–6].

Beyond its cutaneous manifestations, LS carries a substantial burden on patients’ quality of life (QoL). Chronic itching, pain, dyspareunia, architectural changes of the genitalia, and the psychological weight of a visible, often stigmatized condition converge to affect multiple domains of daily functioning[7–9]. Among these, sexual health appears to be one of the most consistently and severely compromised, with patients reporting reduced desire, difficulties with penetration, body image disturbances, and relational strain[10–13]. Testimonies collected from patient communities further underline how sexuality is frequently perceived as one of the most silenced and least addressed aspects of living with LS, despite its central role in overall wellbeing[14].

The compromise of sexual health in LS is best understood as the product of converging physical, psychological, and interpersonal mechanisms rather than a single, purely mechanical consequence of tissue damage. Anatomically, chronic inflammation drives progressive scarring and architectural distortion of the genitalia which can directly impede penetration and diminish sexual function (Figure 1)[15]. Physical symptoms, including pruritus, burning, and pain, sustain a self-reinforcing cycle of anticipatory anxiety and avoidance, in which fear of post-coital flares discourages sexual activity even in patients with comparatively limited objective tissue damage[16]. Moreover, the disease’s relapsing-remitting course, together with frequent diagnostic delay, further fosters hypervigilance toward symptoms and unpredictability that undermines sexual spontaneity[14], underscoring that sexual dysfunction in LS is a genuinely multidimensional phenomenon.

**Fig 1.** Major architectural changes in male (left) and female (right) genital LS. Left panel: typicl LS of the penis with preputial involvement, determining phimosis. Right panel: Clitoral phimosis and labia minora fusion in vulvar LS.

Despite this recognized impact of LS on sexuality, the assessment of sexual dysfunction in LS remains methodologically inconsistent. Several psychometric instruments have been employed across studies, including The Female Sexual Function Index (FSFI)[13] and the International Index of Erectile Function (IIEF)[17], but each carries important limitations when applied to LS. Generic, validated tools such as the FSFI and the IIEF capture core physiological domains of sexuality (desire, arousal/erectile function, orgasm, satisfaction, pain), but are strictly sex-specific, preventing direct, unified comparison between affected men and women and offering limited coverage of disease-related psychosocial dimensions such as body image, partner communication, and avoidance behavior[11,13]. This gap is particularly relevant given emerging evidence that male LS is a clinical entity with its own sexual and functional consequences[17], as shown by recent comparative analyses of sexual health across sexes[5]. Dermatology-specific quality-of-life instruments, including Skindex-29, typically contain at most a single item addressing sexual functioning, which is insufficient to characterize a domain as clinically relevant as sexuality in LS[18].

Against this background, the Qualisex questionnaire has progressively emerged as a reliable, patient-reported outcome measure (PROM) addressing both male and female sexuality within immune-mediated disease, making it a promising candidate for use in chronic inflammatory conditions such as LS, where a validated, sex-inclusive instrument is still lacking. Qualisex is a ten-item self-administered questionnaire, and was originally developed in French to assess the impact of rheumatic diseases on sexual life[19]. Successively, Qualisex has been tested in other languages[20], cultural contexts and medical conditions[21,22] (i.e., in Spain, on people diagnosed with rheumatoid arthritis (RA)[23] and axial spondyloarthritis[24], and in Italy, exclusively on women, who suffered from Sjögren’s syndrome[25] and fibromyalgia[26,27].

In this context, Qualisex was selected for the present study for several converging reasons: it was developed and iteratively validated for chronic immune-mediated disease; its item structure is symmetrical and applicable, without modification, to both men and women, allowing sex-stratified comparisons within a single metric; it is brief and self-administered; it has already demonstrated robust psychometric properties across multiple cultural adaptations.

The aim of the present study was therefore to validate the Italian version of the Qualisex questionnaire for use in patients with LS, providing clinicians and researchers with a reliable tool for the assessment of the disease impact on sexual life in both women and men. Our primary goal was to evaluate the psychometric performance of Qualisex in a sample of patients diagnosed with LS, stratified by sex. In particular, the instrument’s structural construct, validity and reliability were assessed. Discriminant validity against the Depression, Anxiety and Stress Scale-21 (DASS-21) as well as convergent validity against Skindex-29, FSFI and IIEF were also explored.

## Methods

### 2.1 Study Setting and Participants

Data from a cohort of consecutive patients who suffered from LS and considered eligible and agreed to take part to this study were collected between February and July 2026. All procedures involving human participants were in accordance with the Helsinki Declaration. The study protocol was approved by the Local Ethics Committee (Latium Area 5, protocol n. 804/1-IDI). Written informed consent was obtained from all individual participants involved in the study.

The inclusion criteria were (1) patients aged eighteen or above; (2) an established clinical and/or histopathological diagnosis of LS (3); ability to read, write, and understand Italian; (4) willingness to be enrolled in the study and (5) compliance with the study procedures. Underaged patients or patients not willing to participate into the study were excluded. Each participant was asked to fill in the following psychometric instruments: DASS-21, Skindex-29, Qualisex, and IIEF or FSFI (respectively for man or for women).

### 2.2 Administered Questionnaires

The DASS-21 questionnaire is a self-administered tool composed by 21 items rated on a four-point Likert scale (from 0 = did not apply to me at all, to 3 = applied to me very much or most of the time). It measures the negative emotional states indicative of the presence of depression, anxiety and stress (7 questions each)[28]. A General Distress (GD) score is calculated as the sum score of all the 21 items. Higher the score higher the level of emotional distress. The DASS-21 does not have a cut-off score, but it is possible to determine five degrees of severity (from “normal” to “extremely severe”). In this study we used the Italian version of DASS-21 questionnaire[29].

The Skindex-29 is a dermatology-specific Quality of Life (QoL) questionnaire developed originally in English to capture symptoms, emotional impact, and functioning in patients with skin diseases. The questionnaire is self-administered and is composed by three subscales: Symptoms, Emotions, and Functioning[30]. Skindex-29 has been adapted and validated in multiple languages, including Italian[31]. Subscale scores are calculated on a Likert-scales (ranging from 0, ‘never’ to 4, ‘all the time’. For each specific subscale, the numerical scores assigned to the respective items within that domain are summed. The raw scale scores successively undergo a linear transformation into a 0-100 range to enhance interpretability. Higher scores are indicative of a lower quality of life and greater perceived disease severity.

The Qualisex is a self-administered questionnaire; the score is calculated by computing the mean of the results for the 10 items (noteworthy, the likert scale of the last item is purposefully inverted compared to the rest). Higher scores indicate higher disease impact on sexual life. The Italian adaptation and preliminarily validation of Qualisex for sexually active women with primary Sjögren’s syndrome and fibromyalgia has been adopted in the present protocol[25].

The IIEF is widely used in both clinical practice and research settings to assess erectile dysfunction and its severity, and to monitor the efficacy of treatments[32,33]. It was validated in 10 languages while under development and later translated into other 32 languages[34]. The Italian adaptation of the IIEF was used in this study and comprises 15 items evaluating five distinct domains of male sexual health. Each item is scored on a Likert-scale where higher values indicate better function. The “Erectile Function” domain is obtained by summing the first five items and the item #15 (score range: 6–30), which also allows a clinical measure of erectile dysfunction severity (ranging from severe, 6–10, to no dysfunction, 26–30). The remaining subscales include “Orgasmic Function” (items 9 and 10; score range: 0–10), “Sexual Desire” (items 11 and 12; score range: 2-10), “Intercourse Satisfaction” (items 6, 7, and 8; score range: 0–15), and “Overall Satisfaction” (items 13 and 14; score range: 2–10). A global IIEF score ranging from 5 to 75 can be calculated by summing all 15 items, where lower scores represent worse sexual impairment.

The FSFI is considered the gold standard for screening, and one of the most widely used questionnaires to assess the impact of diverse medical conditions and treatments on sexual function in women[35,36]. The Italian adaptation of FSFI used in this study is a 19-item instrument assessing six specific domains of female sexual health. Each subscale score is normalized by applying a specific factor weight to balance its contribution to the final profile. The domains are operationalized as follows: Desire (items 1–2, factor weight 0.6), Arousal (items 3–6, weight 0.3), Lubrification (items 7–10, weight 0.3), Orgasm (items 11–13, weight 0.4), Satisfaction (items 14–16, weight 0.4), and Pain (items 17–19, weight 0.4). Individual weighted domain scores range from 0 to 6. The global FSFI total score, ranging from 2 to 36, is the sum of all six weighted dimensions. An FSFI total score of 26.55 has been found to be the optimal cut-off for differentiating women with and without sexual dysfunction[37].

### 2.3 Statistical Analysis and Psychometric Validation

Two complementary analyses were performed to assess whether the Qualisex effectively captures the true impact of LS on patients’ sexual lives (construct validity). First, Principal Component Analysis (PCA) was used to examine the factor structure of the 10-item instrument. Our goal was to assess if all items merged into a single overarching factor—representing the global burden of the disease on sexuality—or if they split into distinct sub-dimensions (e.g., physical impact versus psychological and relational impairment).

Cronbach’s Alpha coefficients were calculated to evaluate internal consistency of individual items and to assess if they track cohesively together. A threshold of alpha > 0.80 to confirm strong construct reliability was used[38]. Convergent validity with instruments shown to appropriately measure sexual health was assessed using two gender-specific scales (FSFI in women and IIEF in men).

Furthermore, we tested the Qualisex performance against the Skindex-29 in order to assess its clinical utility as a skin-specific tool. We hypothesized that higher Qualisex scores (indicating greater sexual impairment) would scale progressively with poorer dermatological quality of life. This relationship would confirm that the questionnaire successfully captures the unique psychodermatological condition caused by vulvar and penile lesions.

Furthermore, we assessed Qualisex as an independent diagnostic tool, conceptually and empirically separated from general psychological distress, to explore discriminant validity. To test this possibility, we distinguished the specific sexual impact from General Distress (GD). Given that anxiety and depression are frequently comorbid whit chronic dermatological conditions[39–41], we hypothesized a weak-to-moderate correlation (*r* between 0.40 and 0.60) between Qualisex and DASS-21 GD score[42]. A very high correlation (r > 0.90) would therefore indicate construct redundancy, suggesting Qualisex to merely reflect generalized psychological distress. Conversely, a moderate correlation confirms that-despite affective status naturally impacting sexual health-Qualisex captures disease-specific variance that standard, domain-general psychological inventories fail to detect.

Finally, given the hypothesis that Qualisex may possibly reflect more precisely the psychosocial and emotional domains of Skindex-29 subscale, rather than the ones related to physical symptoms, we further looked at how the Qualisex correlated with the specific subscales of the Skindex-29. This pattern could prove mathematically that the Qualisex successfully isolates the relational, interpersonal, and sexual restrictions of the disease from non-specific somatic pain or general physical discomfort, demonstrating a more fine-grained discriminant capacity of such questionnaire.

## Results

### 3.1 Population characteristics

A total of 211 patients with a confirmed diagnosis of LS were enrolled in the study, of whom 167 (79.1%) were women and 44 (20.9%) were men, reflecting the expected female predominance of the condition while still allowing for a sex-stratified analysis of a clinically meaningful male subgroup. The mean age of the overall sample was 52.9 (±11.8) years, with female patients being on average slightly older than male patients (mean 53.8 Vs 47.5 y.o.), consistent with previous reports suggesting an earlier age of clinical presentation or diagnosis in men[43]. Demographic and clinical characteristics of the study population, stratified by sex, are summarized in Table 1.

**Tab 1.**
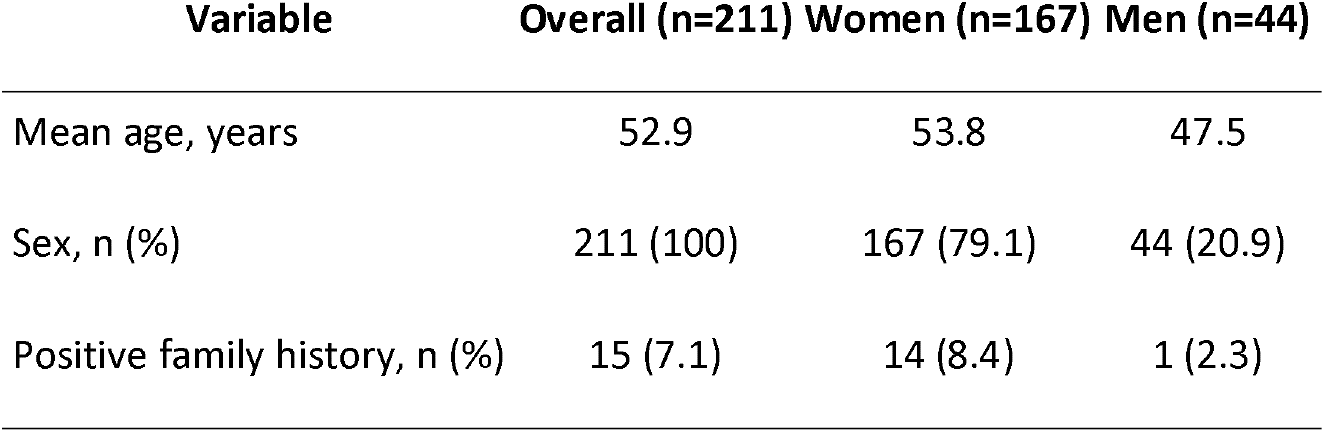
Demographic and clinical characteristics of the study population.

### 3.2 Construct validity

A PCA was performed on the total combined sample. Initially, an oblique Oblimin rotation was applied for enabling the detection of potential inter-factor correlations. As the resulting component correlation matrix yielded a negligible inter-factor correlation (r = 0.079), an orthogonal approach was preferred for parsimony. Consequently, a Varimax rotation with Kaiser normalization was chosen, maximizing the variance of the squared loadings to ensure clear factor separation. Diagnostic sampling adequacy was confirmed by the Kaiser-Meyer-Olkin measure (KMO = 0.932) and Bartlett’s test of sphericity (*χ*^2^(36) = 1285.01, p < 0.001). During initial factor extraction, Item 2 (*“During the past 3 months: have the medications you take been responsible for a worsening of your sex life?”*) demonstrated an unacceptably low communality (0.167). To optimize scale structure, Item 2 was permanently removed. The subsequent PCA on the remaining 9 items yielded a two-factor solution based on the Kaiser criterion (eigenvalues > 1), accounting for 72.5% of the total variance. Component 1 (“Sexual Functioning and Satisfaction”, 59.89% of variance) comprised Items 1, 3, 4, 5, 7, 8, and 9, with strong factor loadings (predominantly > 0.80). This primary dimension captures the biological, somatic, and performance-related impact of Lichen Sclerosus. Component 2 (“Partner Devaluation”, 12.66% of variance) was anchored by Item 6 (*“Have you felt devalued by your partner?”*, loading = 0.895), isolating the interpersonal dynamic from direct physical impairment. Item 10 (*“Overall satisfactory sex life”*) exhibited primary loading on Component 1 (0.591) and a cross-loading on Component 2 (−0.344). To align its direction with the overall scale (where higher scores represent greater impairment), Item 10 was reverse scored. Despite Item 6 displaying a lower corrected item-total correlation (r = 0.258), its retention was theoretically and clinically justified, supporting the calculation of a single global score (Qualisex Total Score). The final 9-item scale demonstrated excellent overall internal consistency (Cronbach’s α= 0.911).

In terms of internal consistency and psychometric performance by gender subsamples, the total 10-item Qualisex scale was highly reliable for females (Cronbach’s alpha = 0.841). Inter-item matrices revealed a relevant closeness between health-driven sexual worsening (Item #1) and both functional performance degradation (Item #4; r = 0.856) and diminished sexual desire (Item #3; r = 0.754). Somatic markers such as pathology-related pain (Item #8) and general fatigue (Item #9) correlated moderately with overall performance impairment (r = 0.661 and r = 0.651, respectively), demonstrating that the psycho-dermatological burden reaches well beyond mere physical pain. Comparatively, external standard validity checks show an appreciable internal consistency for the validation batteries: FSFI total score (alpha = 0.953), DASS-21 total score (alpha = 0.945), and all Skindex-29 subscales (alpha range 0.800-0.880); achieving such high reliability across these established scales strongly suggests adequate respondent attentiveness, rendering a pattern of random or careless responding highly unlikely. This solid internal consistency indicates that the participants filled out the questionnaires with due diligence, thereby reinforcing the overall trustworthiness of the psychometric findings. Similarly, the male sample replicated these strong psychometric features, with the Qualisex total score achieving excellent internal consistency (alpha = 0.899). Corrected item-total correlations remained high (>0.60) for core components, including health perception, performance, pain, and reduced attractiveness. Item #6 (“devaluation”) tracked independently from physical markers (r = 0.019), mirroring the trend observed in the female cohort. Parallel external instruments validated check yield encouraging results: IIEF total score (alpha = 0.964), DASS-21 total score (alpha = 0.963), and Skindex-29 total score (alpha = 0.966).

To test if the Qualisex could purely isolate the psycho-dermatological burden of Lichen Sclerosus rather than age-related physiological declines of sexual functioning, correlation analyses were performed against chronological age (Table 2); while gold-standard functional scales (i.e., FSFI and IIEF) expectedly demonstrated significant negative correlations with age due to natural physiological transitions, the Qualisex total score remained entirely unaffected by age across both sexes (p > 0.05).

**Tab 2.**
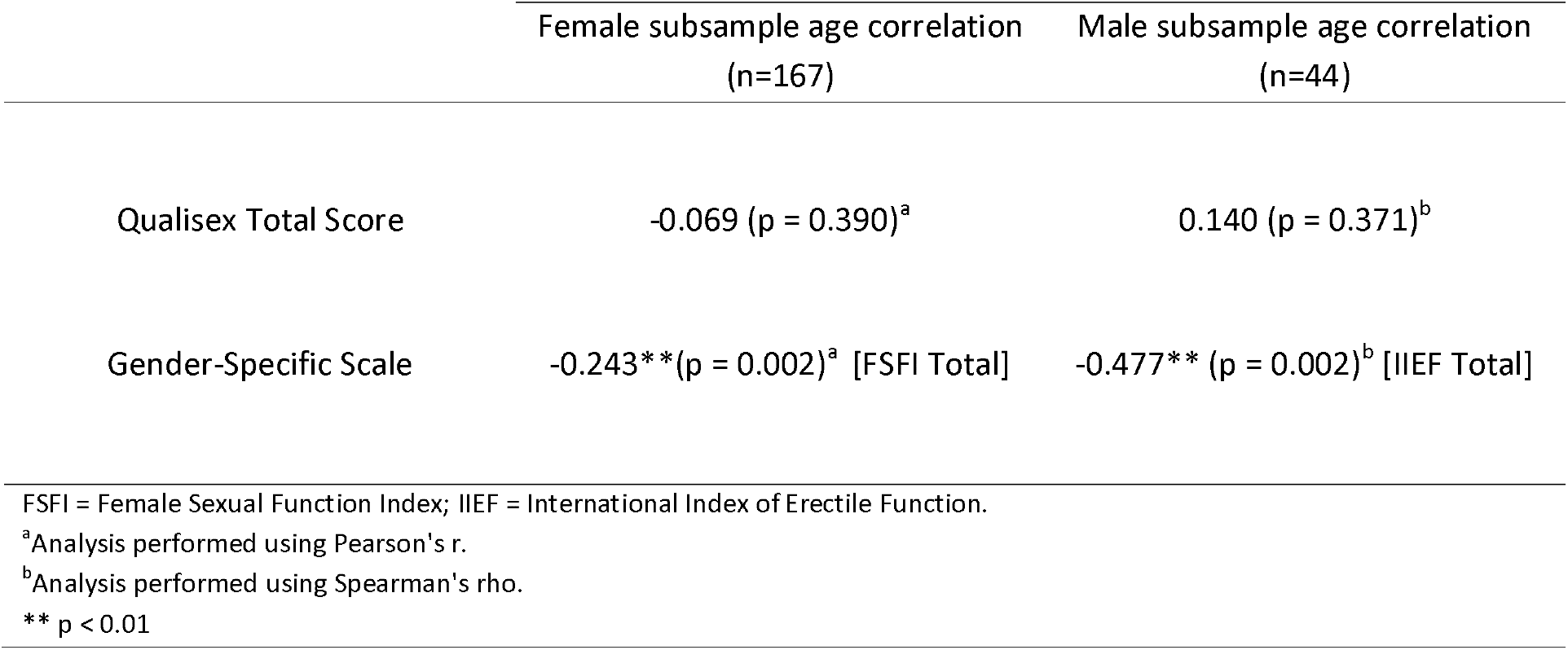
Correlational analysis between age and psychometric measures by genders: divergent validity of the Qualisex Total Score.

The Qualisex Total Score showed a robust convergent validity across both dermatological and sex-functional measures. Statistically significant positive associations were observed with all subscales of the Skindex-29; the strongest associations emerged with the Social Functioning (r = 0.582, p < 0.001) and Emotions (r = 0.531, p < 0.001) dimensions, while a lower, though highly significant, coefficient was found for the physical Symptoms subscale (r = 0.444, p < 0.001). Gender-stratified analyses further confirmed this convergent pattern with established sex-functioning instruments. In the female subscales, higher Qualisex scores significantly correlated with poorer sexual functioning, as indexed by lower FSFI total scores (r = −0.391, p < 0.001). In the male subscale, a strong negative correlation was replicated with the IIEF total score (rho = −0.566, p < 0.001).

Finally, discriminant validity analyses indicated that the Qualisex successfully isolates unique variance directly tied to the specific psycho-dermatological impact of Lichen Sclerosus, without becoming psychometrically redundant, in fact, across both gender cohorts, the Qualisex Total Score showed only a stable, low-to-moderate positive correlation with general psychological distress (DASS-21 Total). Because these coefficients remain below the standard statistical redundancy threshold (r < 0.70), it is possible to state that the instrument does not merely function as a proxy for generic anxiety or depression screeners. This distinctiveness is further highlighted when compared against exclusively sex-functioning tools, particularly within the male subsample. While the IIEF total score did not reach statistical significance when correlated with general psychological distress (DASS-21 Total score: rho = −0.229, p = 0.162) — likely due to the smaller sub-sample size— the Qualisex captured the broader psychological burden of the condition. Taken together, these patterns reasonably show that the Qualisex uniquely operates at the intersection of somatic impairment and emotional well-being, preserving its independence from both pure psychiatric distress and purely mechanical sexual dysfunction scales (Table 3).

**Tab 3.**
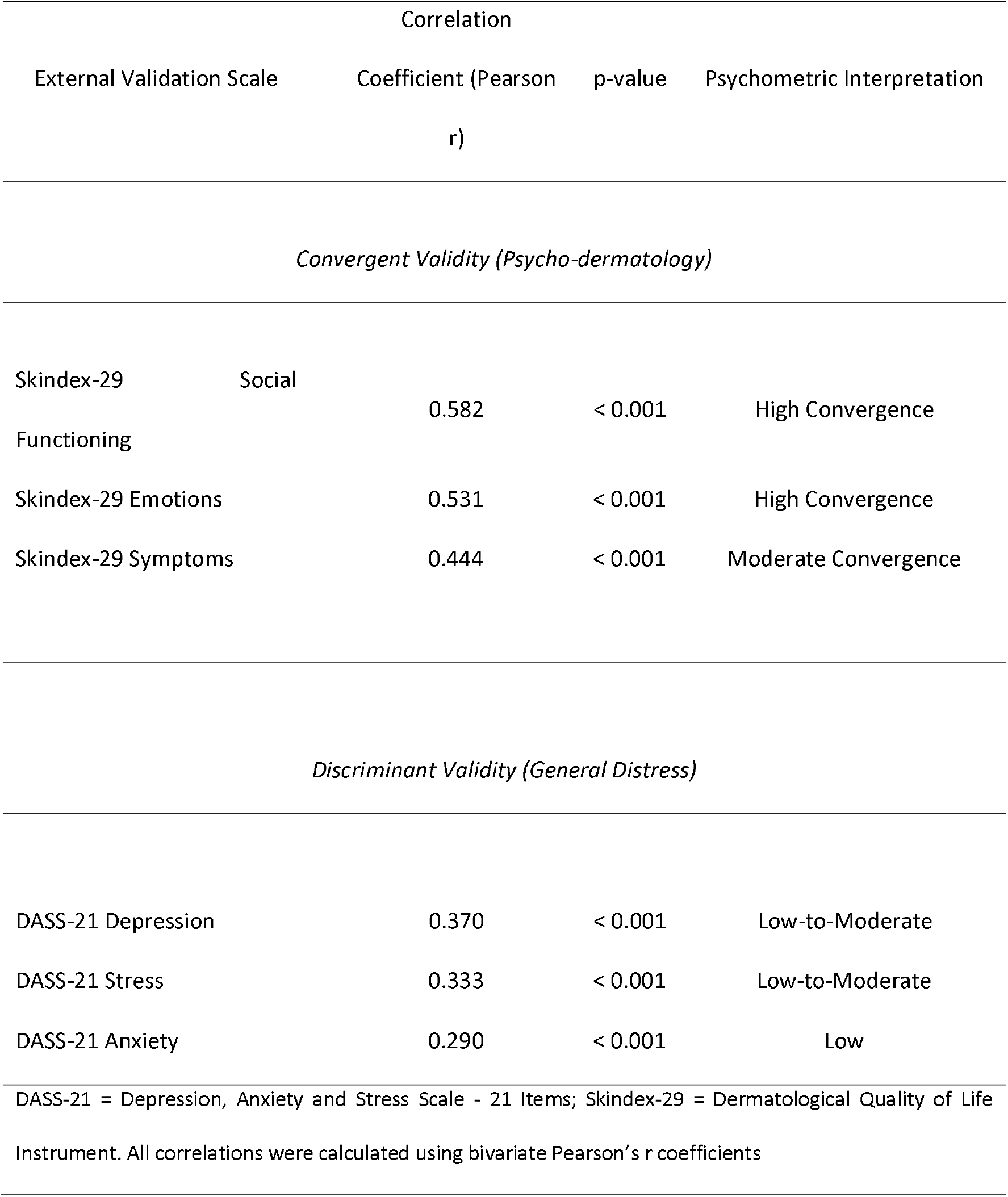
Construct validity matrix of the Qualisex Total score: convergent and discriminant validation gradients (N=211)

## Discussion

The present study evaluated the psychometric performance, component structure, and clinical utility of the Italian adaptation of the Qualisex questionnaire in a sample of patients affected by LS. To our knowledge, this represents the first formal validation of a brief, sex-inclusive PROM specifically designed to capture the psychodermatological burden of LS on sexual health across both male and female populations.

Our findings highlight that the Qualisex questionnaire possesses good psychometric properties, featuring a high internal consistency in LS across the overall scale and both sex subsamples (see Section 3.2). These reliability metrics align closely with previous cross-cultural adaptations in rheumatology, such as the Spanish adaptation for people who suffered from RA (α = 0.971)[23], Turkish adaptation for Axial Spondyloarthritis (α= 0.910)[20,24], and preliminary Italian validations in patients suffering from primary Sjögren’s syndrome (pSS) (α= 0.860), and fibromyalgia (FM) (α= 0.878)[25,27].

However, while most prior adaptations in rheumatic diseases supported a unidimensional, single-factor structure[20,25], our PCA uniquely supported a two-factor architecture ( “Sexual Functioning and Satisfaction” as the primary factor capturing the biological, somatic, and performance-related impact of the disease, and “Partner Devaluation”, uniquely anchored by Item 6 (*“feeling devalued by one’s partner”*). Although this tracked independently from direct physical symptoms, its retention within the global score is both psychometrically and clinically sound. It is important to highlight that LS is well known to trigger deep relational strain, fear of post-coital flares, and genital self-image disturbances[44–46]. Isolating partner devaluation from direct physical dysfunction highlights that sexual distress in LS extends far beyond local mechanical impairment[47].

Item-level refinement further shaped the final scale. It is important to know that item structural adjustments are not unprecedented in Qualisex adaptations; for instance, the Spanish RA study[45] flagged incoherent response patterns on Item #10 due to its reverse-scoring format, and proposed supplementary items. Our initial data extraction led to the permanent removal of Item #2 (“medication-related worsening”), optimizing the structural integrity of the scale without compromising its clinical domain coverage. From a clinical perspective, omitting Item #2 is highly coherent with the therapeutic profile of LS: standard anti-inflammatory regimens—predominantly high-potency topical corticosteroids—are designed to suppress local inflammation and restore mucosal integrity, thus alleviating symptoms rather than directly inducing sexual dysfunction[15].

Given the primarily anogenital localization of LS, sexual health represents a core disease-specific outcome rather than a secondary quality-of-life domain; having a brief, sex-independent, and validated instrument such as Qualisex is therefore particularly valuable, as it allows the psychosexual burden of the disease to be captured using a single standardized metric across both male and female patients, overcoming the comparability limitations of sex-specific tools such as FSFI and IIEF[48].

The major contribution of this study lies in addressing a methodological limitation of previous research. Prior Italian adaptations of Qualisex in pSS and FM explicitly restricted their cohorts to female patients, justifying this design by the female predominance of those rheumatologic conditions[26,27]. In contrast, while LS also exhibits a female predominance, penile and perianal LS in men represent an increasingly recognized and clinically significant entity with distinct functional and psychosexual consequences. By demonstrating strong reliability and convergent validity with the IIEF in male patients, our study establishes Qualisex as a sex-symmetrical tool that enables unified, cross-gender comparative research. Another relevant finding is the age-independence of the Qualisex Total Score. While standard sex-specific questionnaires (FSFI and IIEF) expectedly displayed significant negative correlations with chronological age due to physiological aging processes (Table 2), the Qualisex Total Score did not, remaining stable across age groups. This highlights the instrument’s capacity to effectively isolate the specific psychosexual burden of LS from physiological, age-related sexual decline[49]. Concerning convergent and discriminant validity, the Qualisex Total Score exhibited strong convergent validity across both dermatological quality of life (Skindex-29) and sex-specific (FSFI and IIEF) scales, with the strongest associations emerging for the Social Functioning and Emotions subscales relative to physical Symptoms (Table 3). This gradient of associations seems to mirror findings in pSS, where Qualisex scores correlated with subjective symptoms (e.g., pain, fatigue, dryness) and psychological state rather than objective systemic disease activity markers. Furthermore, discriminant validity analyses suggest that Qualisex captures distinct variance rather than merely reflecting generalized psychological distress, remaining well below the standard redundancy threshold with all DASS-21 domains. While affective distress and anxiety are common comorbidities in chronic genital dermatoses, Qualisex seems to specifically capture the unique intersection between cutaneous pathology, intimate relationships, and sexual well-being.

While the present study provides preliminary evidence for the psychometric properties and clinical utility of the Qualisex scale, several methodological limitations regarding patient selection and cross-cultural generalizability must be acknowledged. First, due to the voluntary nature of recruitment strategy, an a priori power analysis could not be conducted, and the final sample size was determined by the overall response volume within the recruitment window. Second, non-responders or refusal rates were not recorded, precluding a formal assessment of non-response bias. Lastly, while our findings establish a clear two-factor structure in the present sample, future research should evaluate the cross-cultural invariance and factor stability of the scale across different linguistic and clinical contexts.

## Conclusions

The Italian version of the 9-item Qualisex questionnaire is a reliable, validated, and efficient tool for assessing the impact of LS on sexual health. Its brief self-administered format, gender symmetry, and ability to isolate disease-specific impairment from both age-related sexual decline and general psychological distress make it highly suitable for routine clinical consultations and for research. Replicating these findings in independent international cohorts will be essential to confirm whether this factor architecture holds true globally or reflects specific socio-cultural nuances in how sexual impact and partner dynamics are expressed.

## Supporting information

9-item Qualisex

## Data Availability

All data produced in the present study are available upon reasonable request to the authors

## Acknowledgement

The authors sincerely thank Dr. Muriel Ruffaneau and LISCLEA (Italian Lichen Sclerosus Patient Association) for the support in the realization of the present project.

## Statements

### Statement of Ethics

#### Study approval statement

*This study protocol was reviewed and approved by Latium Area 5 Ethics Committee, approval number 465/IDI/26 (804/1-IDI)*

#### Consent to participate statement

Patients involved in this manuscript have given written informed consent to publication of their sociodemographic and clinical data in anonym aggregated form. Informed consent was also obtained for the publication of clinical pictures.

### Author Contributions

Conceptualization: AP, RP; Data curation: LZ, TS, CA; Formal analysis: TS; Investigation: AP, TS, LZ; Methodology: TS; Project administration: AP; Software: TS; Supervision: LC, SG, GDZ; Validation: All Authors; Visualization: All Authors; Writing – original draft: TS and AP; Writing – review & editing: VG. All authors read and approved the final version of the manuscript.

### Data Availability Statement

The data that support the findings of this study are not publicly available due to privacy reasons but are available from the corresponding author upon request.

