## Supplementary material for "Validation of the Qualisex Questionnaire for Psychometric Assessment of Sexual Health in Lichen Sclerosus": 9-item Qualisex

**Supplementart Table 1 S1.**

**Italian validated version of Qualisex questionnaire for lichen sclerosus.**

The Qualisex score is the mean of the results for the 9 questions on a 0–10 scale, with Q10 having a reverse-scoring format. Higher scores indicate more impact of the disease on sexuality. Missing data: if only 1 of the questions is unanswered (or answered as not applicable), the Qualisex score is the mean of the other questions.

Le seguenti domande riguardano l’impatto della sua malattia sulla qualità della vita sessuale. Segni il numero che meglio descrive la sua esperienza negli ultimi 3 mesi.

Q1: Durante gli ultimi 3 mesi: il suo stato di salute è stato responsabile di un peggioramento della sua vita sessuale?

| 0 | 1 | 2 | 3 | 4 | 5 | 6 | 7 | 8 | 9 | 10 |
| --- | --- | --- | --- | --- | --- | --- | --- | --- | --- | --- |

Per niente	Moltissimo

Non mi riguarda []

Q2: Durante gli ultimi 3 mesi: il suo stato di salute ha causato una riduzione del suo desiderio sessuale?

| 0 | 1 | 2 | 3 | 4 | 5 | 6 | 7 | 8 | 9 | 10 |
| --- | --- | --- | --- | --- | --- | --- | --- | --- | --- | --- |

Per niente	Moltissimo

Non mi riguarda []

Q3: Durante gli ultimi 3 mesi: il suo stato di salute ha influenzato negativamente le sue prestazioni sessuali?

| 0 | 1 | 2 | 3 | 4 | 5 | 6 | 7 | 8 | 9 | 10 |
| --- | --- | --- | --- | --- | --- | --- | --- | --- | --- | --- |

Per niente	Moltissimo

Non mi riguarda []

Q4: Durante gli ultimi 3 mesi: il suo stato di salute è stato responsabile di un peggioramento della sua intesa con il partner?

| 0 | 1 | 2 | 3 | 4 | 5 | 6 | 7 | 8 | 9 | 10 |
| --- | --- | --- | --- | --- | --- | --- | --- | --- | --- | --- |

Per niente	Moltissimo

Non mi riguarda []

Q5: Durante gli ultimi 3 mesi: si è sentito/a de valorizzato/a dal suo partner?

| 0 | 1 | 2 | 3 | 4 | 5 | 6 | 7 | 8 | 9 | 10 |
| --- | --- | --- | --- | --- | --- | --- | --- | --- | --- | --- |

Per niente	Moltissimo

Non mi riguarda []

Q6: Durante gli ultimi 3 mesi: il suo stato di salute l’ha fatto/a sentire meno attraente dal punto di vista sessuale?

| 0 | 1 | 2 | 3 | 4 | 5 | 6 | 7 | 8 | 9 | 10 |
| --- | --- | --- | --- | --- | --- | --- | --- | --- | --- | --- |

Per niente	Moltissimo

Non mi riguarda []

Q7: Durante gli ultimi 3 mesi: i dolori legati alla patologia sono responsabili di un peggioramento della sua vita sessuale?

| 0 | 1 | 2 | 3 | 4 | 5 | 6 | 7 | 8 | 9 | 10 |
| --- | --- | --- | --- | --- | --- | --- | --- | --- | --- | --- |

Per niente	Moltissimo

Non mi riguarda []

Q8: Durante gli ultimi 3 mesi: la sua fatica è stata responsabile di un peggioramento della sua vita sessuale?

| 0 | 1 | 2 | 3 | 4 | 5 | 6 | 7 | 8 | 9 | 10 |
| --- | --- | --- | --- | --- | --- | --- | --- | --- | --- | --- |

Per niente	Moltissimo

Non mi riguarda []

Q9: Durante gli ultimi 3 mesi: ha avuto una vita sessuale nel complesso soddisfacente?

| 10 | 9 | 8 | 7 | 6 | 5 | 4 | 3 | 2 | 1 | 0 |
| --- | --- | --- | --- | --- | --- | --- | --- | --- | --- | --- |

Per niente	Moltissimo

Non mi riguarda []
